# Using a physical model and aggregate data from Israel to estimate the current (July 2021) efficacy of the Pfizer-BioNTech vaccine

**DOI:** 10.1101/2021.08.10.21261856

**Authors:** Hilla De-Leon, Francesco Pederiva

## Abstract

From the end of June 2021, the state of Israel, where 60% of the population is vaccinated with an mRNA BNT162b2 vaccine, has an increase in the daily morbidity. This increase may be a result of different events: a temporal decline of the vaccine’s efficacy; Lower efficacy of the vaccine against the current Delta ((B.1.617.2) variant (which is now the dominant strain in Israel); A result of lack of social restrictions, a highly contagious variant, or any combination of the above. We found, by using a novel spatial-dynamic model and recent aggregate data from Israel, that this new surge of cases is partiality due to a decline in the shielding of those who were vaccinated about six months ago. Also, we found a decrease in the vaccine’s efficacy against severe morbidity for the early elderly population compared to the rest of the vaccinated population. These results, which are consistent with recent studies, emphasize the high ability of the model in evaluating the time- and age- dependent efficacy of the vaccine for different age groups and enables to predict the spread of the pandemic as a function of such efficacy.

## I. INTRODUCTION

In December 2020, the State of Israel began an extensive vaccination campaign after the FDA granted emergency authorization approvals (EUA) to two mRNA vaccines - the Pfizer-BioNTech and Moderna vaccines. To date, nearly 6 million Israelis are vaccinated with a Pfizer-BioNTech vaccine (at least with the first dose). [1–3]. Hence, many studies have been released that use the epidemiological information currently collected in Israel to examine the vaccine’s success rate compared to the official data provided by Pfizer. The empirical data accumulated on the first month after the beginning of the vaccine operation in Israel indicated the high effectiveness in preventing moderate to severe morbidity and death, especially among patients aged 60 and over, who were the first in Israel to receive the vaccine. Based on the information gathered, it can be estimated with high confidence that the vaccine has an efficacy of at least 90% in preventing morbidity (after 28 days from the first dose) and at least 40% in preventing the infection (see, for example Refs. [3–6]). However, starting from June 2021, the number of confirmed cases in Israel, which has been declining since January 2021, is raising again. There can be many reasons for the increase in the number of confirmed cases, including the removal of most of the social restrictions that have been practiced in Israel since March 2020, on the possibility that the vaccine is less effective against various variants of virus (see, for example, Refs. [7, 8]), in particularly with the current Delta variant [9], the declining in the effectiveness of the vaccine might be a function of time, age-dependency of the efficacy of the vaccine or any combination of the above. In this work, we use both real-time Israeli aggregated data and a diffusive Monte-Carlo (MC) model as introduced by De-Leon and Pederiva (see Refs. [10, 11] and Methods) to predict the current efficacy of the Pfizer-BioNTech vaccine against both morbidly and infection. The strength of this model is that, in contrast to other infection models, such as Susceptible Infected Removed (SIR) [12–14], this model is a particle model capable to distinguish between different age groups and to treat each of them separately, under the assumption that infection occurs in the entire population at the same time. In addition, a particle model allows the adjusting of the rate of population immunization in order to accurately examine the different effects of the vaccine on both subgroups of the vaccinated population and the whole population. This is a straightway to estimate the efficacy of the vaccine.

## II. USING MC DIFFUSIVE MODEL AND REAL DATA FOR PREDICTING THE CURRENT EFFICACY OF THE PFIZER-BIONTECH VACCINE

We use atomistic-like computer simulations to model the spread of the epidemics. In all simulations, we employed a MC diffusive model as introduced by De-Leon and Pederiva (Refs. [10, 11] and methods) to study the expected effects of vaccinations on the evolution of both the confirmed cases (CC) and severe hospitalizations and to estimate the current efficacy of the vaccine. [15]. Based on the information accumulated in Israel over the last year, we found that severe and moderate hospitalization takes place about five days following a positive RT-PCR test (see Ref. [6]). Also, based on the current data from Israel, we found that there is a 5-days shift from being hospitalized to becoming a severe case (Fig. 1), where the number of the new daily severe cases is approximately 0.6 of the new hospitalized five days earlier (all the data was taken from Refs. [1, 2]).

**FIG. 1:**
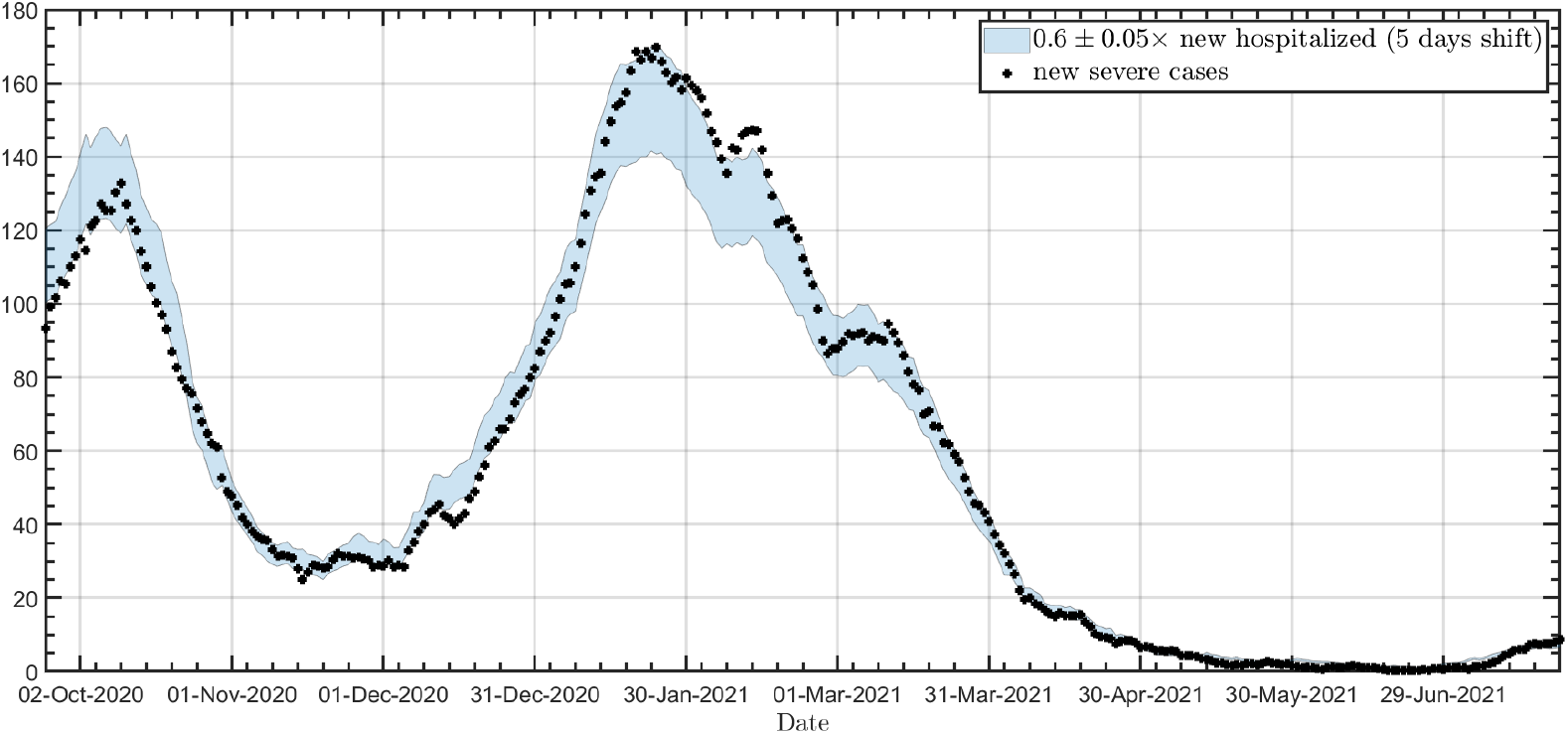
The correlation between the new daily severe and moderate hospitalized and the new severe hospitalized after five days. Dots: new severe hospitalized; Solid blue band: 0.6 ± 0.05× the new severe and moderate hospitalized with five days shift (all the data was taken from Refs. [1, 2]

As the numbers of new severe cases are independent of either testing policy, they consist a clear and robust indication for the dynamics of disease and the efficacy of the vaccines (with a ten-day shift) in contrast to the number of new confirmed cases. Nevertheless, since the data analysis requires a division by age that is accessible only for confirmed and new severe cases, as opposed to Ref. [6], in this work, we evaluate the vaccine’s efficacy based on these data and the model.

The data accumulated so far indicate that the Pfizer-Biontech COVID-19 vaccine has high efficacy against the infection and reduces the transmission probability (Refs.[3, 4]). This high efficacy might allow for a relatively broad opening of the economy without causing a further outbreak once a significant percentage of the population will be vaccinated.

From January 2021 till June 2021, the number of the daily confirmed cases is constantly declining. However, as of June 2021, it is on the rise again as well as the number of daily new hospitalization. In this paper, we will use the combination of the model as introduced in Refs. [10, 11] and the aggregated data available from Refs. [1, 2] to estimate the current efficacy of the vaccine and to predict to spread of the pandemic in Israel giving that efficacy.

Estimating the efficacy of the vaccines against both infection and severe morbidly is required to distinguish between *R*_*t*_ (the theoretical effective reproduction number of the virus) and *R*_*e*_ (the effective reproduction number of the virus). *R*_*t*_ estimates the number of encounters between carriers and healthy people who would have ended in infection without the vaccines and define the population’s dynamics level, wheres *R*_*e*_ is affected by the vaccination rate and by the vaccine’s protection against infection (see Section B in methods for more details).

For this work, based on the removal of social restrictions in Israel starting in February 2021, we assumed that the theoretical *R*_*t*_ in Israel is increasing over time (see Methods). However, till June 2021, this increase in *R*_*t*_ did not reflected in an increase in Israel’s confirmed cases. In Israel, the first persons to be vaccinated were the citizens over the age of 60. This broad vaccination operation was the cause of the decline in the number of new confirmed and hospitalized cases from mid-January 2021, especially for people above 60 [6, 16]. Fig. Ref. 2 shows the percentage of the confirmed cases for 60 and up from all the confirmed cases from December 2020, as well as the total daily confirmed cases. From Fig. 2 we find that the current rise in the confirmed cases in Israel is accompanied by a higher increase in the confirmed case of the age of 60 and over, in contrast to the past six months. This increase may indicate a current lower efficacy of the vaccine compared to January.

**FIG. 2:**
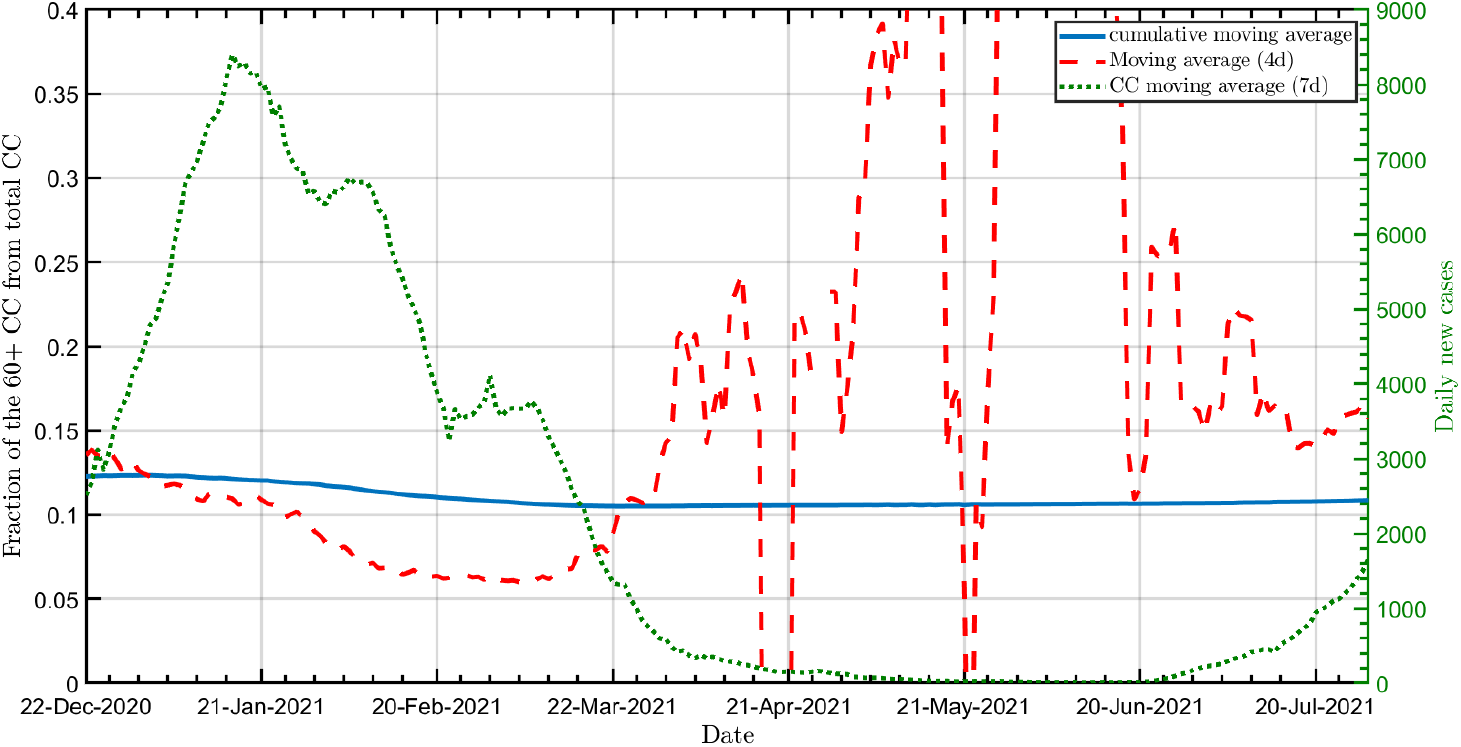
Percentage of verified persons aged 60+ among all verified persons as of January 2021. Solid line: cumulative moving average dashed line: 4 days moving. Dots: the daily total confirmed cases.(all the data was taken from Refs. [2**?**]

Figure 3 shows our prediction for the spread of the pandemic in Israel since January 2021 for four different efficacies, where all the models assume the same *R*_*t*_ and the current vaccination rate in Israel (see Methods). We showing in Fig. 3 our prediction for the spread of the pandemic for three age groups: (a) The total population (b) The elder population (age 60 and up), which can use as a measure to estimate whether there is a decline of the vaccine’s efficacy over time. (c) The young population (under 16), which can use as a measure of the ability of the vaccine to protect even those who are not covered by a vaccine [17] (currently, about 90% of 0-16-year-old in Israel are not protected by the vaccine [1, 2]). Note that for both Figs. 3 and 4, to avoid numerical artifacts, we split our prediction into two parts in which for each part, the model was calibrated separately; a. from January 2021 till May 2021 (third wave in Israel). b. from the end of June 2021 (fourth wave in Israel).

**FIG. 3:**
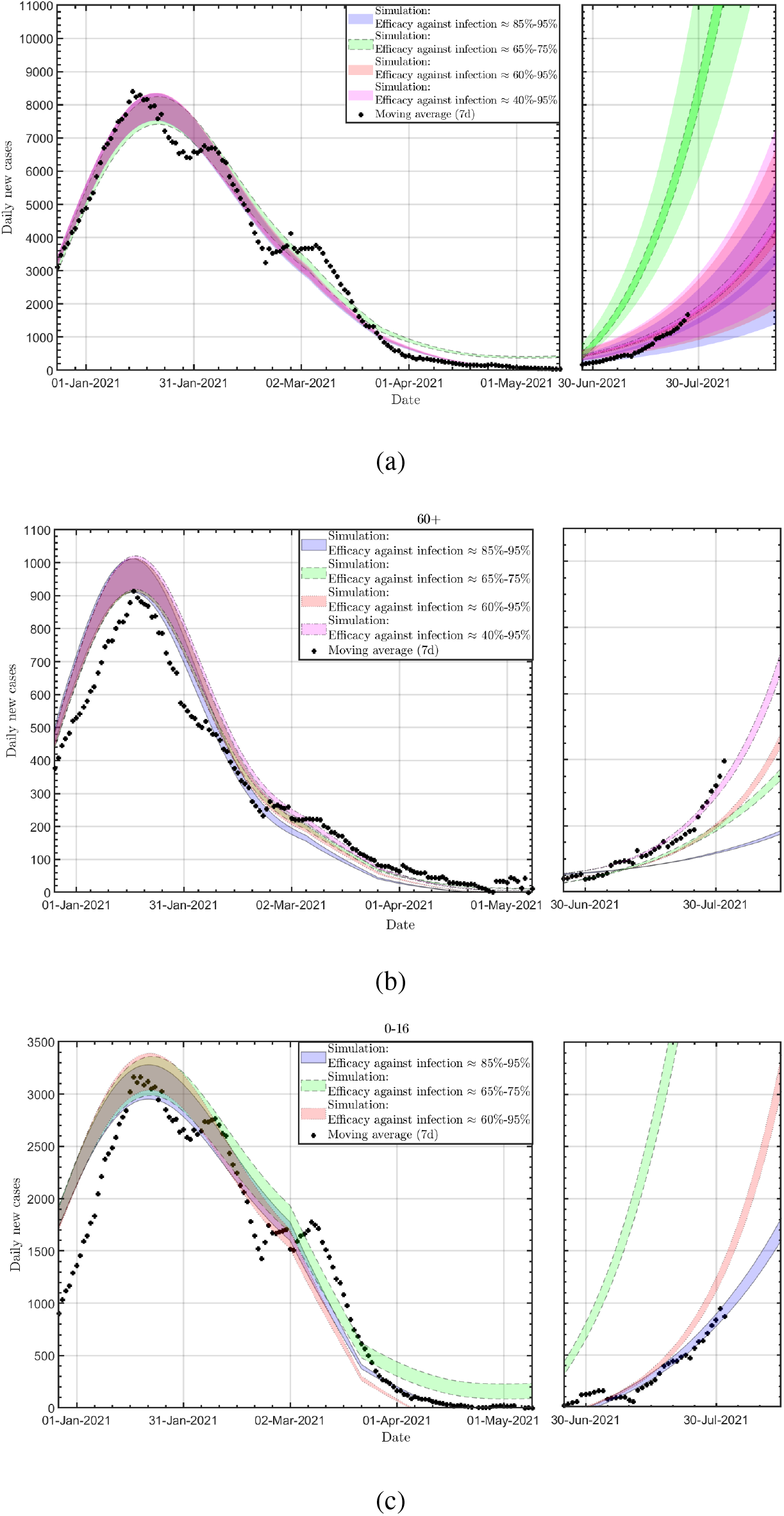
The daily new confirmed cases since January 2021 of the total population (a); people over 60 (b) and people under 16 (c). For all panels: Solid blue band: 90% efficacy against infection; Dashed green band: 70% efficacy against infection; Dotted red band: 90% efficacy against infection for the first 150 days after the second dose of the vaccine and 60% efficacy against infection after 150 days since the second dose of the vaccine; Dotted-dashed magenta band: 90% efficacy against infection for the first 150 days after the second dose of the vaccine and 60% (40%) efficacy against infection after 150 days since the second dose of the vaccine for people under (above) the age of 60. Crosses: Real data (7-day moving average). The shaded areas in panel (a) represent the uncertainly that propagated from that on *R*_*t*_. The uncertainty in panels (b) and (c) is similar to that of panel (a) and is omitted for improving the readability of the figure.

**FIG. 4:**
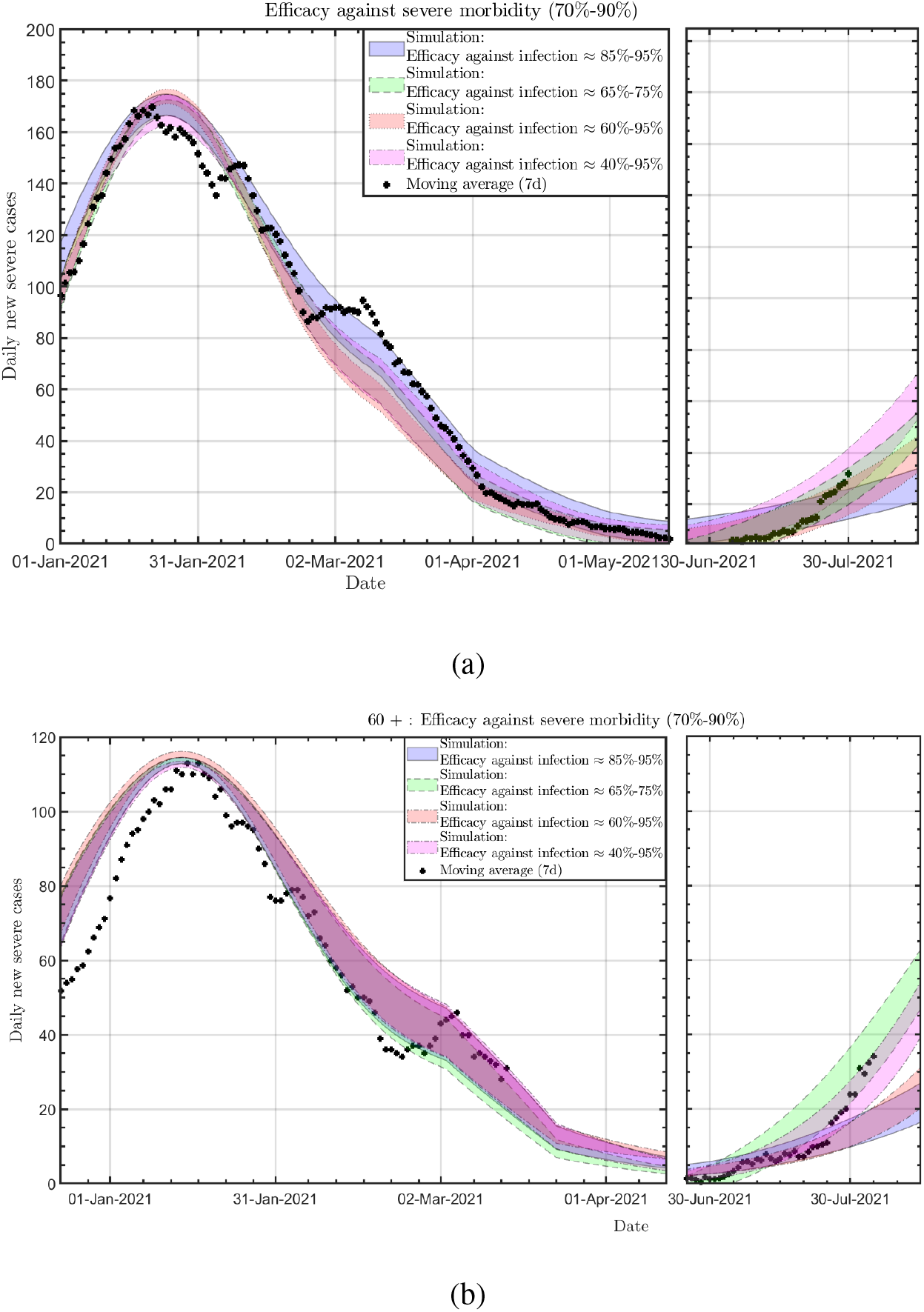
The daily new severe cases form January 2021 of the total population (a) and of people over 60 (b). For both panels: Solid blue band: 90% efficacy against infection; Dashed green band: 70% efficacy against infection; Dotted red band: 90% efficacy against infection for the first 150 days after the second dose of the vaccine and 60% efficacy against infection after 150 days since the second dose of the vaccine; Dotted-dashed magenta band: 90% efficacy against infection for the first 150 days after the second dose of the vaccine and 60% (40%) efficacy against infection after 150 days since the second dose of the vaccine for people under (above) the age of 60. Crosses: Real data (7-day moving average) Crosses: real data (7-days moving average).

From Fig. 3 one finds that for the case of uniform efficacy (solid blue band and green dash band in Fig. 3 there is no correlation in terms of confirmed cases between the total number of confirmed cases and the number of confirmed cases of people above the age of 60. We found that while the number of confirmed cases of the young population is consistent with a model assuming high efficacy against infection (solid blue band), the number of confirmed cases at older ages is not compatible with this efficacy and implies a lower efficacy (dashed green band).

Therefore, based on the high percentage of the confirmed cases over 60 (the first vaccinators) from the total confirmed cases, for the third model, we assume high efficacy (90%) in the first five months after the second dose of the vaccine. Then the efficacy decreases to 60% efficacy against infection (dotted red band in Fig. 3). Also, we tested a fourth model in which we assume a high efficacy (90%) in the first five months after the second dose of the vaccine. Then the efficacy decreases to 40% (60%) against infection for the over (under) 60 population. From Fig. 3 is shown that the third and fourth models manage to predict the number of overall and the young confirmed cases as well as the number of confirmed cases aged 60 plus. (Note that since the model uses a limited number of particles for predicting morbidity, and based on a comparison between the model and morbidity in Israel in August-December 2020, we assume a 10% error of the model).

Another stringent test of the reliability of the model is its ability to predict the total daily new severe cases and of those above 60 as shown in Fig. 4. Similar to Ref. [6], we assume that the chance of hospitalization is a function of age and workload in the hospital, with suitability for patients in severe condition. Similar to our prediction of the confirmed cases, we examined four different efficiencies against severe morbidity. For all four models, we assume the same vaccine efficacy against infection as presented in Fig. 3. Giving the estimation of 90% against severe morbidity, for the first three models, we assume an 85% - 95% efficacy against severe morbidity (for all ages), which is reflected in the width of the different bands in Figure 4. For the fourth model, we assumed that for the elderly population, the vaccine’s efficacy against severe morbidly declines after five months to 60%-70% for the elderly population.

Note that we realize that today, the information of the new daily severe cases is limited and makes it difficult to fit the data to a specific model. However, we use this data as an additional test for the model, knowing that shortly, we can better estimate the vaccine’s effectiveness in preventing severe morbidity.

Nevertheless, for daily new severe cases, similar to Fig. 3, both real data and the model indicates that there is a time-dependent declined in the efficacy of the vaccine against infection in comparison to its efficacy at the beginning of 2021, while the efficacy of the vaccine against severe morbidly is still relatively high.

Also, when examining both Fig. 3 and Fig. 4 we found that, based on our estimation of *R*_*t*_ between January and May 2021, it is hard to distinguish between the two (constant) efficiencies in terms of the continuing decline in the number of daily new confirmed cases and the daily severe cases. However, with the current increase in the number of daily confirmed cases, one finds out that the vaccine’s efficacy is a significant factor in daily morbidity. Hence, since the rate of morbidity is a function of both *R*_*t*_ and vaccine efficacy (as shown in the upper panel of Fig. 3), we use the maximum reported efficacy of the vaccine to assess the upper limit of *R*_*t*_ from January 2021 till today (see section M.III in Method), where, in principle, one can assume a lower *R*_*t*_ and lower efficacy of the vaccine for the same morbidity. However, considering a lower *R*_*t*_ will require us to take very low efficacy for early vaccinated adults, which is inconsistent with current studies [18, 19].

## III. CONCLUSION

In this paper, we used aggregated data from Israel and a dynamic Monte Carlo algorithm to estimate the current efficacy of the Pfizer-BioNTech vaccine (ageist the Delta, B.1.617.2, variant) using four different scenarios of vaccine efficacies. The model’s main advantage is its extreme flexibility, such that we can test many efficacy scenarios for different age groups with a minimal computational cost.

This paper examined how a time- and age-dependent decline in the vaccine’s efficacy can affect the pandemic outbreak using a theoretical model and aggregate data from Israel. Our model indicates that while the vaccine’s effectiveness against infection of most of the population is still high (85% − 95%,, which is consistent with Ref. [19]), we detect a decrease in vaccine’s protection against infection for the population vaccinated more than half a year ago. These results are similar to the findings of Ref. [18], which found (based on a studied population of more than 1.3 million people) that the risk for infection against the current Delta variant was significantly higher for people receiving an early vaccination compared to those vaccinated later. Also, our current finding indicates a decrease in the vaccine’s efficacy against severe morbidity for early elderly vaccinators.

Hence, this work demonstrates how by using an accurate model, one can detect the vaccine’s efficacy by using **only** aggregated data. Therefore, this model can be used as a tool for predicting the spread of the virus for a given vaccination rate and can use for determining the minimum range of social restrictions that will prevent the further spread of the pandemic. In particular, this model is relevant nowadays when the elderly population in Israel gets a third dose of the vaccine, which aimed to raise their protection against infection and severe morbidity level to the same level that was in early 2021 [20].

## Data Availability

All the data was taken from:
https://github.com/dancarmoz/israel_moh_covid_dashboard_data
https://data.gov.il/dataset/covid-19

https://data.gov.il/dataset/covid-19

## ACKNOWLEDGMENTS

We thank D. Gazit, Y.Ashkenazy and R. Calderon-Margalit of the Hebrew University COVID-19 pandemic monitoring team for fruitful discussions. HDL thanks Eldad Sitbon for fruitful discussions

## METHODS

We employed models simulating confirmed cases and new moderate and severe cases.

### M. THE MODEL

The simulations in this work use the previously described model ([10, 11]), which models the spread of the pandemic as an interaction between particles in a specific 2-D surface. The simulation performed, assuming an initial surface area unit of four km^2^, and 1.1 · 10^4^ particles which represent the population as a set of interacting individuals, employing the basic mechanism of statistical physics, such that each entity is tagged with four health statuses (susceptible, infected, recovered/dead or vaccinated). In the simulation, a susceptible person can become infected become sick with a daily probability from interaction with all the sick people on the surface. Still, a sick person stops being sick only as a function of time and not as in interactions with a healthy person. In addition, throughout the simulation, a certain percentage of the population began to be vaccinated and, as a result, be protected from infection by the virus. The model’s algorithm is based on standard Monte Carlo (MC) procedures of sampling the transition among the following states, sampled from a statistical distribution, in the spirit of transport MC algorithms. The model used here (similarly to what was reported in Refs. [10, 11]) is a “one-way” Ising-model Monte Carlo such that a healthy person (i) can become sick with a daily probability *P*_*i*_ = ∑ _*j*_ *P*_*i j*_, where *P*_*i*_ *j* is a function of the distance between each infected person (j) in the area and the healthy person (i).

#### M.I. The model’s basic mechanism and infection probability

##### 1. List of parameters, and corresponding values, that describe the population and the kinetics of the infection

For modeling the spread of the pandemic, based on Ref. [21], this work has identified a list of parameters and corresponding values that describe the population and the infection’s kinetics[22].

1. The number of effective individuals, denoted by *N*.
2. The fraction of “latent carriers,” which have no symptoms (AKA asymptomatic) but can infect other people is denoted by *a*_latent_, and the probability of transmitting the infection has been set to 0.5.
3. Each sick person is considered contagious between the 3^rd^ and the 7^th^ day.
4. Simulations are started with a single infected person (the *zero patient*). Also, every day there is a chance of 20% that a healthy person (who is not recovered/vaccinated) becomes sick without interaction with a known sick person.

All the simulations are started with a single infected person (zero patient).

##### 2. Population dynamics

The model is based on the principles of Brownian motion, such that for each day, the population position and displacement (Δ*R*) are given by:

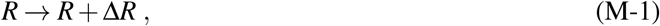

where 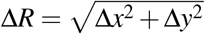 is distributed normally:

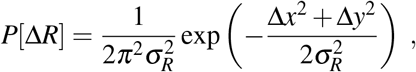

where Δ*x* (Δ*y*) is displacement in the x-(y-) direction 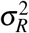, the variance, is a function of the diffusion constant, *D*:

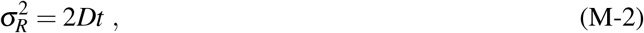

where t=1 day. For a Brownian motion the diffusion coefficient, *D*, would be related to the temperature, *T*, using the Einstein relation:

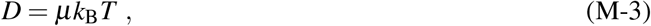

where *µ* is defined as the mobility, *k*_B_ is Boltzmann’s constant and *T* is the absolute temperature. By fixing *T* = 1, the diffusion coefficient would be directly related to mobility. In our model, during all the time period, the population is allowed to move with *σ*_*R*_ = 500 meter.

##### 3. The infection probability

The core of the model, that contains most of the epidemiological data, is the probability for the i^*t*^*h* healthy person to become sick. We assume that for each contact with another sick person this process can be described by a Gaussian function of the distance, weighted with a factor that parametrizes the conditions and social interaction of the sick person:

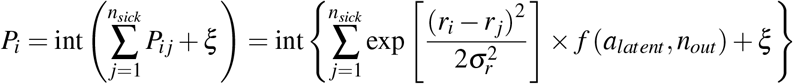

where:

- int is the integer part
- *r*_*i*_ (*x*_*i*_, *y*_*i*_) is the location of the *i*^th^ healthy person and *r* _*j*_ (*x* _*j*_, *y* _*j*_) is the location of the *j*^th^ sick person, so |*r*_*i*_ − *r* _*j*_| is the distance between them.
- *n*_*sick*_ is the total number of sick people in the area.
- *σ*_*r*_ is the standard deviation (here *σ*_*r*_ = 2.4 meters) since recent studies show that even a slight breeze can drive droplets arising from a human cough over more than 6 meters [23].
- *f* (*a*_*textlatent*_, *n*_*out*_) is a function that considers the social activity of the sick person and whether he has symptoms, which affects the spread of the virus outside the house. In our model, we estimate that infection by asymptomatic people is approximately 50% lower than patients with symptoms.
- *ξ* is a random number uniformly distributed between 0 and 1, needed for converting to infection probability to a number between 0 and 1.
- We are assuming that each sick person will infect some of his household members. Since the latest estimates are that household infections are ∼ 15% from the known cases (without lockdown, [24]), we estimated that the number of household infections is uniformly distributed between 0 and 3.
- During all the runs, we assume that people must wear face-masks so that the daily infection probability (for the non-household members, eq. M-4) is reduced to [25–29]

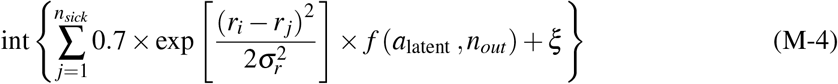

- For all the simulations, people are forced to maintain a distance of 3 meters from each other (social distancing).

#### M.II. Adding vaccination by age groups to the model

All simulations simulate the morbidity situation in Israel since the beginning of December when on December 21, the State of Israel began vaccinating the entire population over 16. The first to be vaccinated were those aged 60+ followed by those aged 50+ and so on (see Fig. S-1). Therefore, the model includes a division by the age of the verified cases as follows: The simulation consists of 11,000 particles, where each particle has a number from 1 (youngest) to 11,000 (oldest). Since in Israel 17% of the patients were above the age of 60, we assumed that the highest 17% numbers in the model represent the population over the age of 60 in Israel, which were the first to be vaccinated in the simulation (and so on to all the age groups) (the daily vaccination rate was taken from Refs. [1, 2].

Using the model, we can analyze the spread of the epidemic and its morbidity in Israel by examining the vaccine’s efficiencies in preventing infection and morbidity and comparing it to Israel’s actual data. In all the simulations, we assumed some protection against infection, which ranges from 40% to 95 % after 21 from the date vaccination. Also, we assumed that vaccinated individuals’ infectivity is 50% (infected vaccinated patients are less infected than unvaccinated patients). In addition, we examined, using the model, the vaccine’s protection against severe morbidity as a function of the time elapsed since the vaccination (see Section C). The visual comparison between the model and the actual data from Israel for both confirmed cases and severe and moderate (SM) morbidity enabled us to evaluate (with 10% uncertainty) the vaccine’s effectiveness against infection and morbidity.

#### M.III. Real time dynamic

For the model, the important parameters are just the transition probabilities, vaccine effectiveness and rate of population vaccination [1, 2], as well as the effective reproduction number of the virus,*R*_*e*_. The daily *R*_*e*_ that is used as an input to the simulation is the weekly averaged 4-day growth factor in Israel, until the onset of the third lockdown (January 8, 2021). This is done using the relation between *R*_*e*_ and the particle density (i.e., the area where the simulation occurs, denoted by *S*) such that:

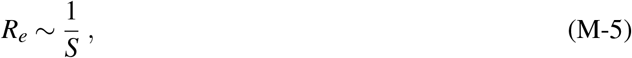

since *R*_*e*_ represents the number of potential encounters in the population, the area reduction is equivalent to increasing the population density and consequently increasing *R*_*e*_.

**FIG. M.1:**
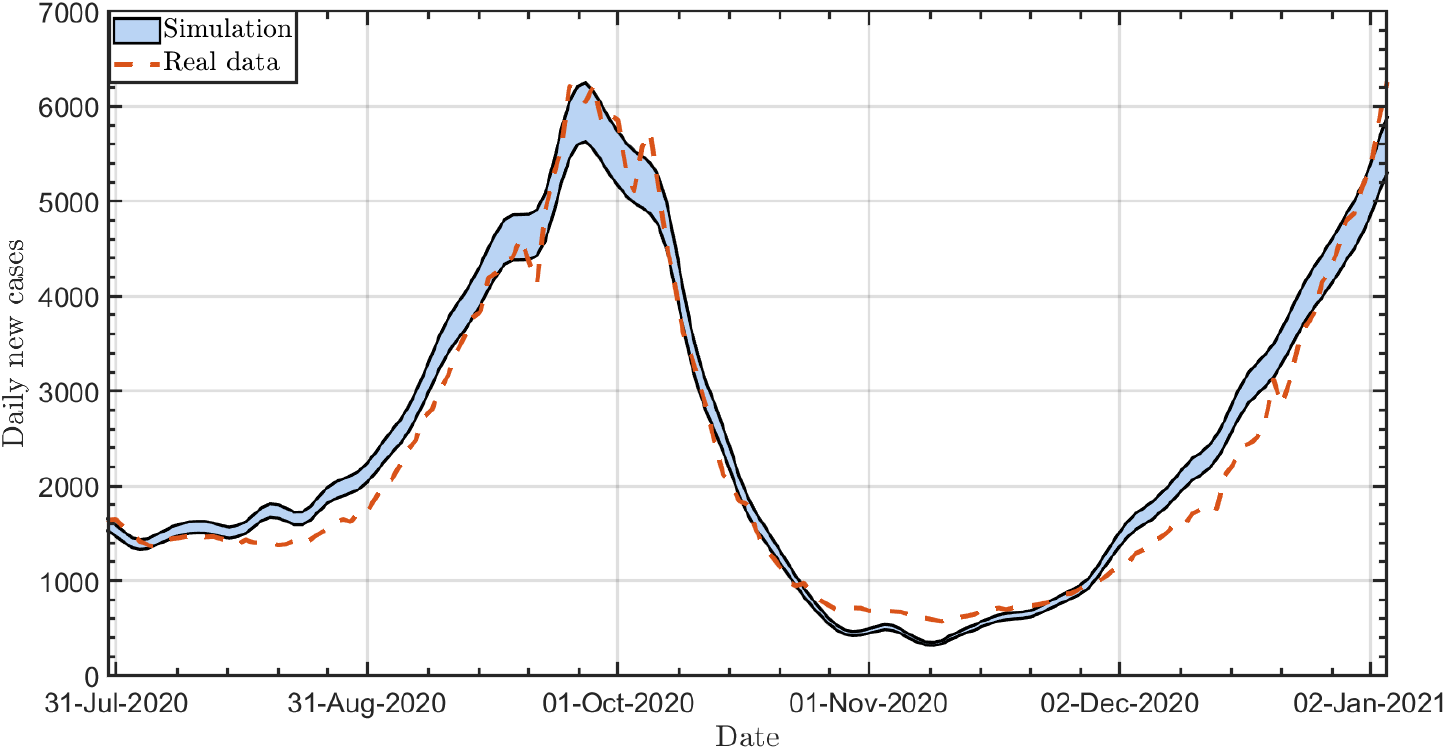
The daily new confirmed cases between Aug 1st and Jan 6th, 2021. Solid line: simulation. Dashed line: real data [1, 2]

Figure M.1 shows the agreement of the simulated daily new cases to the reported data in Israel from August 1, 2020, till January 6, 2021, based on the effective reproduction number, *R*_*e*_ in Israel. This bolsters the model’s validity and enables us to use it in this work for future predictions.

However, given the vaccination campaign in Israel, a distinction is required between *R*_*t*_, the theoretical effective reproduction number of the virus, and *R*_*e*_ (effective reproduction number of the virus). *R*_*t*_ represents the number of encounters between carriers and healthy people who would have ended in infection without the vaccines and defines the population’s dynamics level, where *R*_*e*_ is affected by the vaccination rate and the vaccine’s efficacy against infection.

Although it is not trivial to estimate *R*_*t*_, we can assume that for the total population:

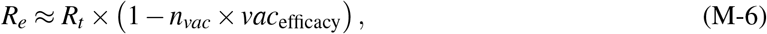

where: *n*_*vac*_ is the percentage of vaccinated in the population and *vac*_efficacy_ is the efficacy of the vaccine against infection.

Therefore, since *n*_*vac*_ is known and both the model and the data are age dependant, the correct combination of *R*_*t*_ and *vac*_efficacy_ should restore not only the total number of confirmed cases but also the total number of confirmed cases for each age group separately.

From January 8, 2021, until February 18, 2021, when the Israeli government imposed a national lock-down, the population compliance is simulated such that the population behavior corresponds to *R*_*t*_ = 1.1, i.e., each carrier meets on average 1.1 people that would have been infected without vaccines. From February 18 until today, we used In the product: 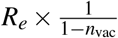 as an upper limit for our estimation of *R*_*t*_.

**FIG. M.2:**
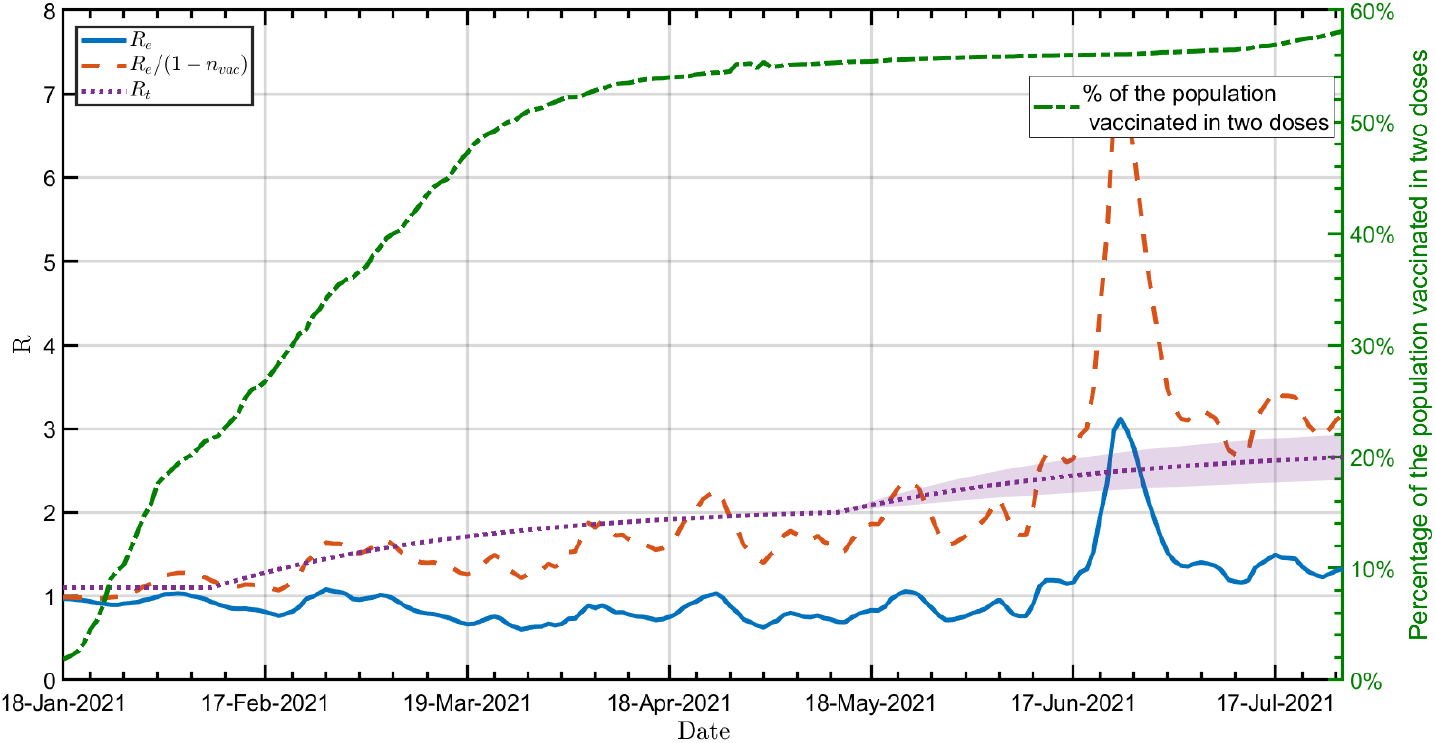
The confirmed cases effective reproduction number of the virus, *R*_*e*_, as a function of time. Sold line: *R*_*e*_, based on the daily confirmed cases in Israel; Dashed line: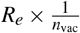;Dotted line: *R*_*t*_ =1.1 at lockdown and has been rising gradually since then with 10% uncertainty form the begging of June (shade area); Dotted-dashed line: the percentage of the Israeli population vaccinated in two doses [1, 2]

Figure M.2 shows the real effective reproduction number, *R*_*e*_, (based on the daily confirmed cases in Israel), which is a result of both the population behavior (*R*_*t*_) and the effect of vaccines (solid line).

#### M.IV. Moderate and severe daily admissions as an indicator for the state of the pandemic in Israel

Based on previous work [6], we use moderate and severe daily admissions as a measure for the disease in Israel. This is justified by the fact that there exists a correlation between this parameter and the population infection rates. This is exemplified in Fig. M.3, which showed that there is an agreement between the effective reproduction number estimated by the 4-day growth factor of either detected cases (full line), with its standard deviation (shaded area), or using the moderate and severe new daily hospitalizations (dashed line). Maximal correlation is achieved by shifting back the Re of the newly confiremd cases by five days. As confirmed cases are delayed by five days on average from infection in Israel, this is consistent with a 10-day average deterioration time from infection to moderate or severe condition. As a result, this parameter is a clear and robust indicator for a COVID-19 severe case that has the advantage of being evident quite promptly after infection but does not depend on detection or population test compliance. Thus, it is ideal as an objective measure of disease dynamics and the effect of the vaccine on the outbreak.

**FIG. M.3:**
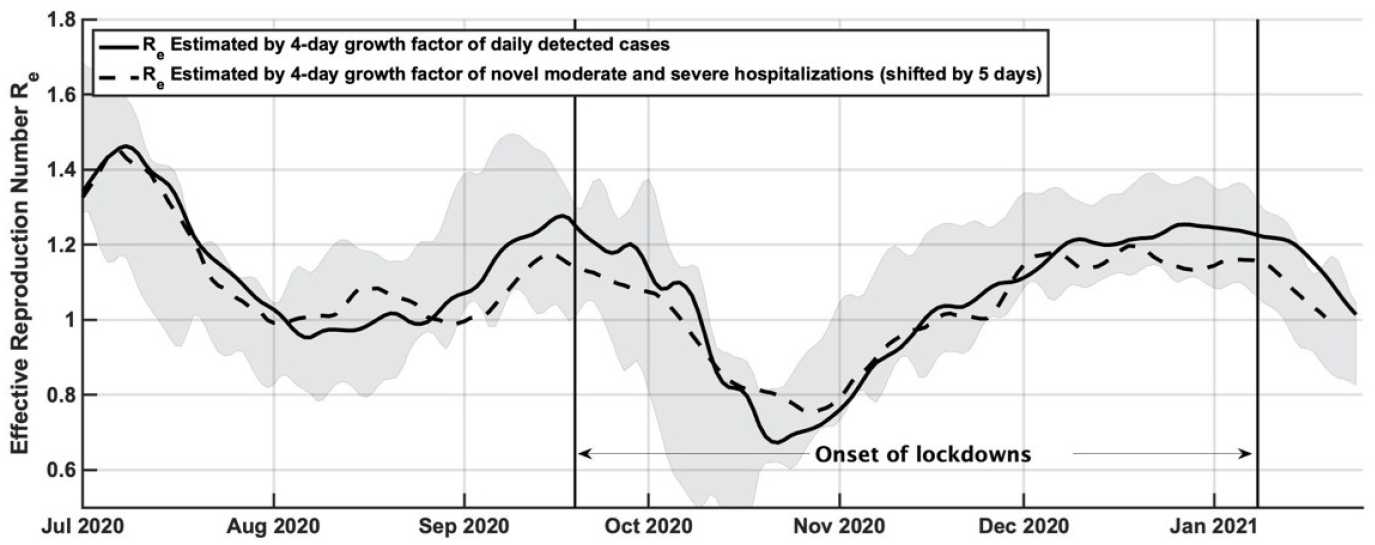
Solid line: Effective reproduction number estimated by the 4-day growth factor of either detected cases (full line), with its standard deviation (shaded area); Dashed line: moderate and severe new daily hospitalizations, shifted back 5 days to maximally correlate to the detected cases.

#### M.V. Simulating the time dependence of new moderate and severe cases and age distribution

The inputs *R*_*e*_ and *R*_*t*_ allow a prediction of the number of confirmed cases. As shown by using Fig. M.3, this leads to admissions in moderate or severe condition (and as a result, the number of new severe cases as shown in Fig. 1). We then calibrated the simulation to reproduce the new moderate and severe cases around the second lockdown period (i.e., between August 1, 2020, and October 31, 2020). The calibration process leads to a fit of the probability of a person being hospitalized in this condition. It is found that the probability of hospitalization decreases as the load on hospitals increases. This is a measure of the effect of hospital capacity on the treatment an individual receives. Fig. M.4 presents the percentage of moderate and severe morbidity as a function of the daily confirmed cases (with a five-day shift) for four age groups from 19 and up (where the groups were distinguished as mentioned in Section A). Data and their fitted analytical functions are presented for elderly (over 60) and younger (under 60) patients, the severe/moderate morbidity in the current wave can be estimated for both age groups. These functions are then used to predict such morbidity and death.

**FIG. M.4:**
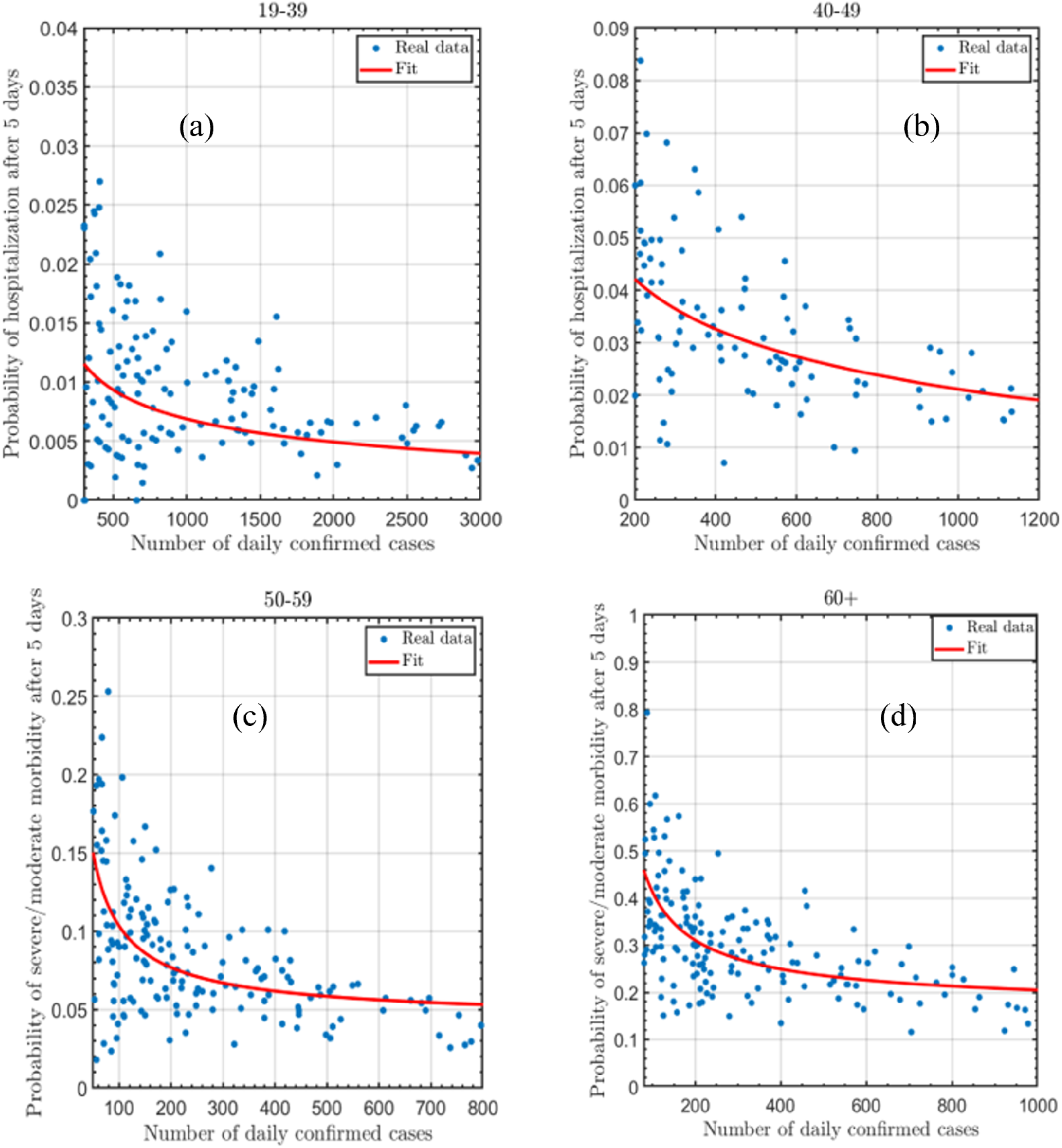
the probability of moderate and severe morbidity as a function of the daily confirmed cases (with a five-day shift) for four age groups; (a):19-39;(b) 40-49;(c) 50-59 (d) 60+. For all panels, the dots represent real data of the moderate and severe morbidity in Israel between 01/08/2020 and 31/10/2020. The solid line is fitted function

